# Clinical referral text overrides MRI evidence in multimodal large language models: glioma versus brain abscess

**DOI:** 10.64898/2026.10.17.26365809

**Authors:** Nedim C. Beste, Fabian Bauer, Nicolás Lopez, Thomas Schömig, Michael Schönfeld, David Zopfs, Lukas Goertz, Thorsten Persigehl, Christoph Kabbasch, Marc Schlamann, David Maintz, Roman J. Gertz, Nils Weitzer, Jonathan Kottlors

**Author notes:** These authors contributed equally (Nils Weitzer, Jonathan Kottlors). **Corresponding author** Nils Weitzer, Institute for Diagnostic and Interventional Radiology, University Hospital Cologne, Kerpener Straße 62, 50937 Cologne, Germany.

## Abstract

**Objectives:** To determine how strongly clinical referral text, rather than image content, drives the decisions of multimodal large language models (LLMs) differentiating glioma from brain abscess on MRI.

**Materials and methods:** This retrospective single-center study included 60 contrast-enhanced T1-weighted brain MRI examinations (30 gliomas, 30 abscesses; reference-standard diagnosis). One axial slice per case was paired with five referral texts (explicitly inflammatory to explicitly tumor-suggestive) under three prompt instructions and submitted 20 times each to GPT-4o, Claude Sonnet 5, Gemini 3.5 Flash, and Qwen 3.6 (72,000 answers). The primary outcome was the text-driven span (difference in tumor-call frequency between explicit inflammatory and explicit tumor text). Two blinded radiologists classified the images without text; sensitivity, specificity, and accuracy were compared descriptively.

**Results:** Patients (29 women) had a mean age of 55.8 ± 16.4 years. With neutral text, models answered tumor in 45–67% of queries (specificity, 0.38–0.78), versus 52% and 60% for radiologists (specificity, 0.77 and 0.63). The text-driven span was 0.79–1.00 (*p* < 0.001 for all models), whereas confirmed gliomas and abscesses differed by only 0.01–0.10 in three models and 0.25 in Gemini 3.5 Flash. Subtle wording produced at least 83% of the span in three models. An instruction to ignore the text reduced the span to 0.48–0.70 in three models and to 0.04 in Gemini 3.5 Flash, partly through reversion to a tumor default. Model accuracy was 0.52–0.73 versus 0.78 and 0.73 for radiologists.

**Conclusion:** Multimodal LLMs deferred to referral text over imaging evidence in differentiating glioma from brain abscess, and prompt instructions did not reliably correct this.

**Key Points:** *Question:* Multimodal large language models receive images together with clinical referral text, but how strongly the text rather than the image determines their diagnosis is unknown.

*Findings:* Changing only the referral text largely reversed the glioma-versus-abscess decision for identical MRI images in four models; instructions to ignore the text reduced this inconsistently.

*Clinical relevance:* Before clinical use, multimodal large language models require model- and task-specific testing of their susceptibility to referral text, because instructing them to prioritize the image is not a dependable safeguard against text-driven misdiagnosis.

## Introduction

A radiologist almost never reads an image on its own. Imaging requests arrive with clinical text (the referral question, symptoms, and laboratory findings), and weighing that text against the image is part of every diagnosis. Multimodal large language models (LLMs) now process image and text within the same system [1, 2] and have been evaluated for report structuring, protocol selection, differential diagnosis, and image-based report generation [3–7]. As these systems move toward clinical use, they will receive images together with the same kind of text that radiologists receive. How they weight the two is therefore a safety question: a model that follows the text against the image can misclassify a lesion in either direction, with the risk of unnecessary or delayed treatment [8, 9].

Prior evaluations have mostly measured diagnostic accuracy on isolated inputs [1–4], and high sensitivity with low specificity has been reported when these models read images directly [2]. Recent work showed that deliberately biased prompts, such as authority claims or anchoring statements, reduce LLM accuracy on radiology board-style questions, most strongly on multimodal items [10]. Whether ordinary, non-adversarial clinical text has the same effect on real cases, how much wording it takes, and how models behave before any suggestive text is added has not been examined.

We chose glioma versus brain abscess as the test case. Both can appear as ring-enhancing lesions on contrast-enhanced T1-weighted imaging, yet they require entirely different treatment, and diffusion-weighted imaging is typically needed to separate them [11–13].

Withholding it keeps the image ambiguous but not uninformative, which is the condition under which the weighting of text against image becomes measurable. An ambiguous image should produce uncertainty, not deference to the text.

The aim of this study was to determine how strongly clinical referral text, relative to the image, determines the decisions of multimodal LLMs. We hypothesized that suggestive clinical text would shift model decisions away from the image, that simple prompt instructions would reduce this effect inconsistently across models, and that models would be as consistent when wrong as when right.

## Materials and methods

### Study design and patients

The ethics committee of the University of Cologne reviewed this retrospective single-center study (reference number 24-1252-retro) and raised no objections; under state data-protection law, the requirement for informed consent was waived. Cases were identified in the radiology information system and picture archiving and communication system (DeepUnity 2.0.2.2; Dedalus HealthCare) of the Institute for Diagnostic and Interventional Radiology, University Hospital Cologne, and selected randomly among examinations acquired between 2014 and 2026. Clinical data were retrieved from the electronic health record of the hospital information system (PORB 08044403.00020.DACHL; Dedalus HealthCare). The reference standard was histopathologic confirmation for gliomas and microbiologic confirmation and/or the documented clinical course for abscesses.

Inclusion criteria were a confirmed reference diagnosis; MRI at 3 T (MR 7700, Ingenia 3.0; Philips) including a thin-section (1 mm) contrast-enhanced T1-weighted sequence acquired after intravenous administration of 15 ml contrast agent (Clariscan 0.5 mmol/ml; GE Healthcare Buchler GmbH & Co. KG); a clearly ring-enhancing intra-axial lesion; and sufficient image quality without relevant artifacts. Exclusion criteria were an equivocal reference standard, relevant image artifacts, and prior surgery or radiotherapy in the lesion area. One representative axial slice per case was used. Diffusion-weighted imaging and all other sequences were withheld by design, so that the image could not force an answer. Images were de-identified, and no protected health information was transmitted to any model. The study design is summarized in Fig. 1. Reporting followed the Checklist for Artificial Intelligence in Medical Imaging (CLAIM) [14].

**Fig 1.**
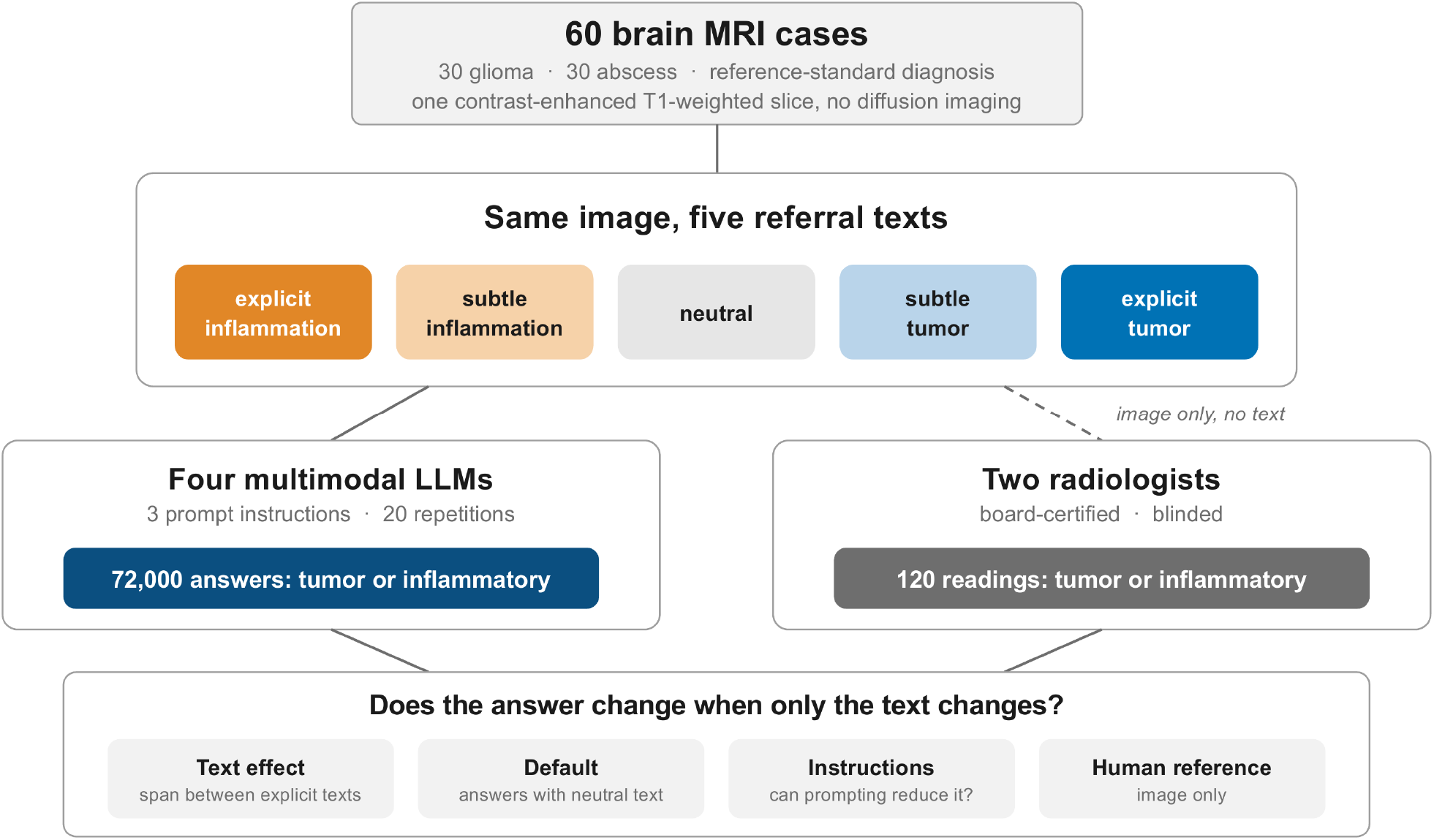
Study flowchart and design. Sixty brain MRI examinations with reference-standard diagnoses (30 glioma, 30 abscess) were presented as one axial contrast-enhanced T1-weighted slice each; diffusion-weighted imaging was withheld. Four multimodal large language models (LLMs) classified each image paired with five standardized referral texts under three prompt instructions, 20 times per condition (72,000 answers). Two blinded board-certified radiologists classified the same images without text (120 readings)

### Clinical text conditions

For each case, five standardized referral texts of the kind that accompanies an imaging request were written for this study and arranged along a bipolar axis: explicit inflammatory, subtle inflammatory, neutral, subtle tumor, and explicit tumor (Supplementary Table 1). The texts and instructions were developed by three radiologists in iterative rounds of revision before the main queries. Patient age, sex, and presenting symptoms were identical across levels, so that only the diagnostic suggestiveness changed; the neutral text was non-directive. Two ablations were added: the image without text, and the neutral text without an image. All prompts were administered in German, the working language of the study site; English translations are provided in the Supplementary Information.

**Table 1.**
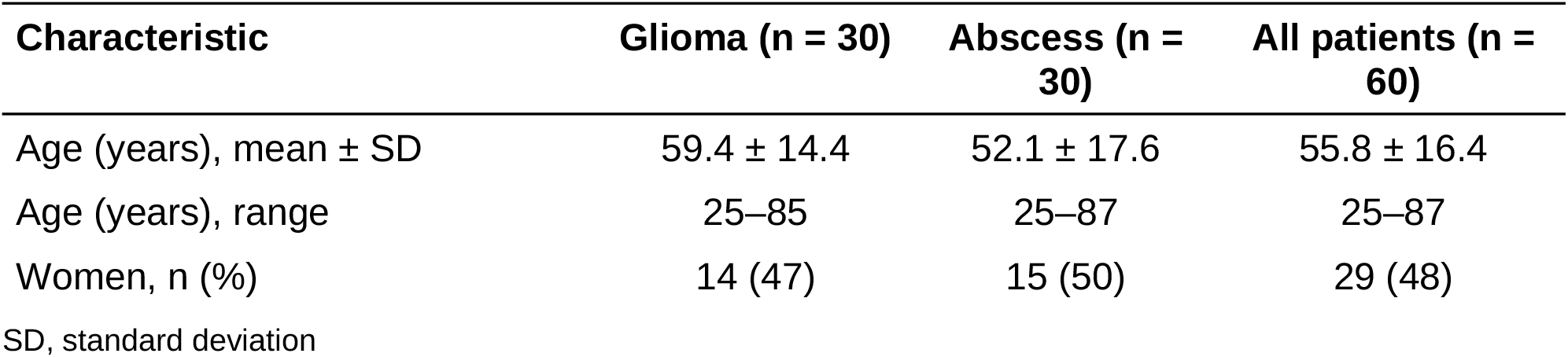
Patient characteristics.

| Characteristic | Glioma (n = 30) | Abscess (n = 30) | All patients (n = 60) |
| --- | --- | --- | --- |
| Age (years), mean ± SD | 59.4 ± 14.4 | 52.1 ± 17.6 | 55.8 ± 16.4 |
| Age (years), range | 25–85 | 25–87 | 25–87 |
| Women, n (%) | 14 (47) | 15 (50) | 29 (48) |

SD, standard deviation

### Models and querying

Four multimodal LLMs were accessed through their application programming interfaces: GPT-4o (gpt-4o; OpenAI), Claude Sonnet 5 (claude-sonnet-5; Anthropic), Gemini 3.5 Flash (gemini-3.5-flash; Google), and Qwen 3.6 (qwen3.6-27b; Alibaba, accessed through Groq), hereafter GPT-4o, Claude, Gemini, and Qwen. All queries were run between May 2026 and August 2026. Each query was an independent, stateless request. Models were asked for a forced choice between tumor and inflammatory, with no option to abstain (Supplementary Table 2). Each condition was repeated 20 times, and only the final categorical label was retained; responses that matched neither category were classified as unclear and excluded. Temperature was set to 0.7 for GPT-4o, Gemini, and Qwen; for Claude, the provider default was used because temperature could not be configured. Provider-default top-p settings were retained. The maximum output length was 20 tokens for GPT-4o and Claude, 500 tokens for Gemini, and 800 tokens for Qwen, to accommodate model-internal reasoning tokens.

### Prompt instructions

To test whether the effect of the text could be controlled, every condition was run under three system-prompt instructions: no weighting instruction, a soft instruction to prioritize the image over the text, and a hard instruction to ignore the text and decide from the image alone (Supplementary Table 2). Instructions were phrased qualitatively rather than as numeric weights, because an LLM does not compute an explicit weighted combination from such a value.

### Human reader reference

Two board-certified radiologists (readers NR1 and NR2), one with completed neuroradiology subspecialty training, independently classified the identical single-slice inputs as tumor or abscess, without clinical text. Readers were blinded to the reference standard and to each other, and the 60 cases were presented in random order. The readings served as an image-only reference for how much diagnostic information the restricted images contained.

### Outcomes and statistical analysis

The primary outcome was the tumor-call frequency, the proportion of the 20 repetitions of a condition in which a model answered tumor. This proportion carries no diagnostic claim by itself; it measures how far the answer moves when only the text changes. For each case and instruction, the text-driven span was defined as the difference in tumor-call frequency between the explicit inflammatory and the explicit tumor text for the same image, averaged across the 60 cases. The share of the span reached with the subtle text was calculated separately for each direction. Secondary measures were the baseline leaning (tumor-call frequency with neutral text) and internal consistency (absolute deviation of the tumor-call frequency from 0.5), compared between correct and incorrect majority answers.

Mixed-effects logistic regression with a random intercept per case modeled the tumor call as a function of text level, instruction, and their interaction; pairwise contrasts used Holm correction. Proportions are reported with Wilson 95% confidence intervals (CIs). Reader performance against the reference standard was summarized as accuracy, sensitivity, specificity, and Cohen’s κ; agreement between the two readers was also assessed with Cohen’s κ. Because the reference comprised only two readers, models and readers were compared descriptively. Analyses were performed in R, version 4.2.1 (R Foundation for Statistical Computing); two-sided *p* values < 0.05 were considered to indicate statistical significance.

## Results

### Study cohort

The final cohort comprised 30 patients with glioma and 30 patients with cerebral abscess (Table 1). Mean patient age was 55.8 years (SD, 16.4; range, 25 to 87), and 29 of 60 patients (48%) were women (glioma, 59.4 years and 14 women; abscess, 52.1 years and 15 women).

### Human reader reference and answers with neutral text

All models returned an interpretable answer in nearly every query; unclear responses occurred in 0% (Claude and Qwen), 0.03% (GPT-4o), and 0.1% (Gemini) and were excluded. With neutral text, before any suggestive text was introduced, the models answered tumor in 67% (Claude), 59% (Qwen), 54% (GPT-4o), and 45% (Gemini) of queries for a balanced case set (Fig. 2a). Specificity for inflammatory lesions was 0.38 (Claude), 0.53 (Qwen), 0.48 (GPT-4o), and 0.78 (Gemini), and sensitivity for tumor was 0.57 to 0.71. The two radiologists, reading the same images without text, answered tumor in 52% and 60%, with specificity of 0.77 and 0.63. Model accuracy was 0.52 (GPT-4o), 0.54 (Claude), 0.62 (Qwen), and 0.73 (Gemini), as compared with 0.78 and 0.73 for the radiologists (κ with the reference standard, 0.57 and 0.47) (Fig. 2b and Supplementary Table 3). The two readers agreed in 45 of 60 cases (75%; κ, 0.50; 95% CI, 0.28 to 0.72). Accuracy is reported only to show that the images were difficult but informative, not as a claim about diagnostic performance.

**Fig 2.**
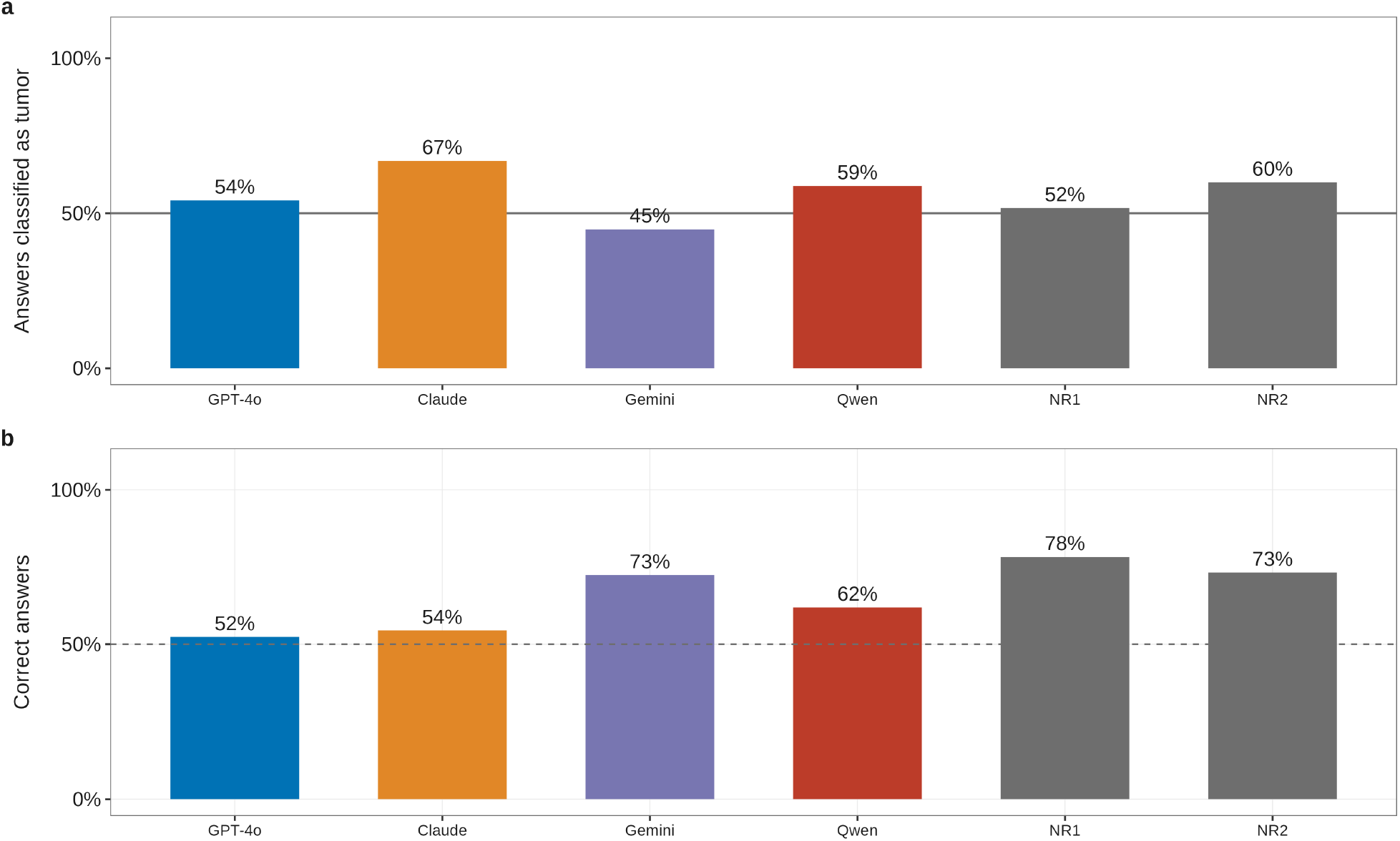
Answers with neutral text and human reference. a Proportion of answers classified as tumor for each model when the text carried no suggestion, alongside two blinded radiologists (NR1, NR2) who read the same images without text. With 30 tumor and 30 inflammatory cases, 50% (solid line) would be balanced. b Proportion of correct answers; the dashed line marks chance level. Accuracy is shown only as context, because a single non-discriminative sequence is not intended to support claims about diagnostic performance

### Effect of the clinical text conditions

Adding suggestive text to the unchanged images moved the answer across almost its entire range. The text-driven span was 1.00 (SE, 0.00) for GPT-4o, 0.97 (SE, 0.02) for Claude, 0.86 (SE, 0.03) for Qwen, and 0.79 (SE, 0.04) for Gemini (*p* < 0.001 for all models). The same image was therefore called tumor or inflammatory depending only on the accompanying text (Fig. 3 and Supplementary Fig. 1). For GPT-4o, Claude, and Qwen, the confirmed diagnosis had little bearing on this: answers for confirmed tumors and confirmed abscesses differed by 0.01, 0.03, and 0.10 on average (maximum, 0.24), far less than the text moved them. Gemini showed a somewhat larger difference between confirmed tumors and abscesses (mean, 0.25; maximum, 0.46), although the effect of the text remained larger (Supplementary Figs. 2 and 3).

**Fig 3.**
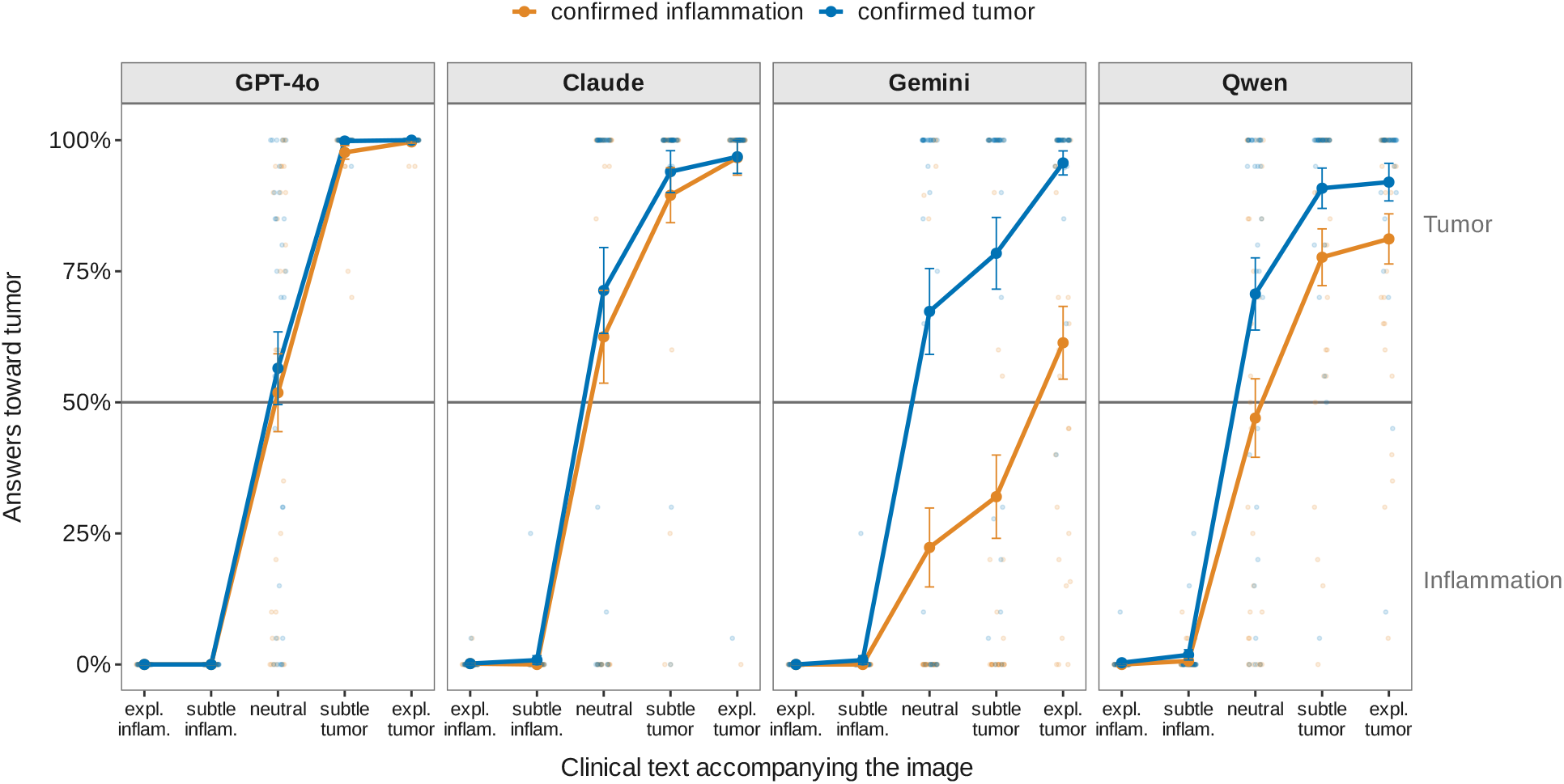
Effect of the text, separated by confirmed diagnosis. The horizontal axis moves in five steps from an explicitly inflammatory to an explicitly tumor-suggestive text; the image is identical at every point, so any change reflects the text alone. The two lines in each panel show confirmed tumor and confirmed inflammatory cases. A model relying on the image would keep the lines apart; in GPT-4o and Claude, they run almost on top of each other (mean difference, 0.01 and 0.03), whereas they are somewhat further apart in Qwen and Gemini (0.10 and 0.25). The horizontal line at 50% marks complete uncertainty

### Subtle versus explicit text

On the tumor side, the subtle text alone produced 98% (GPT-4o), 83% (Claude), and 92% (Qwen) of the change achieved with the explicit text; on the inflammatory side, the corresponding values were 100%, 100%, and 98%. Gemini showed a graded response on the tumor side (31%), whereas on the inflammatory side its subtle text produced 99% of the change (Fig. 4). In three of four models, a faint hint was treated almost like an explicit statement.

**Fig 4.**
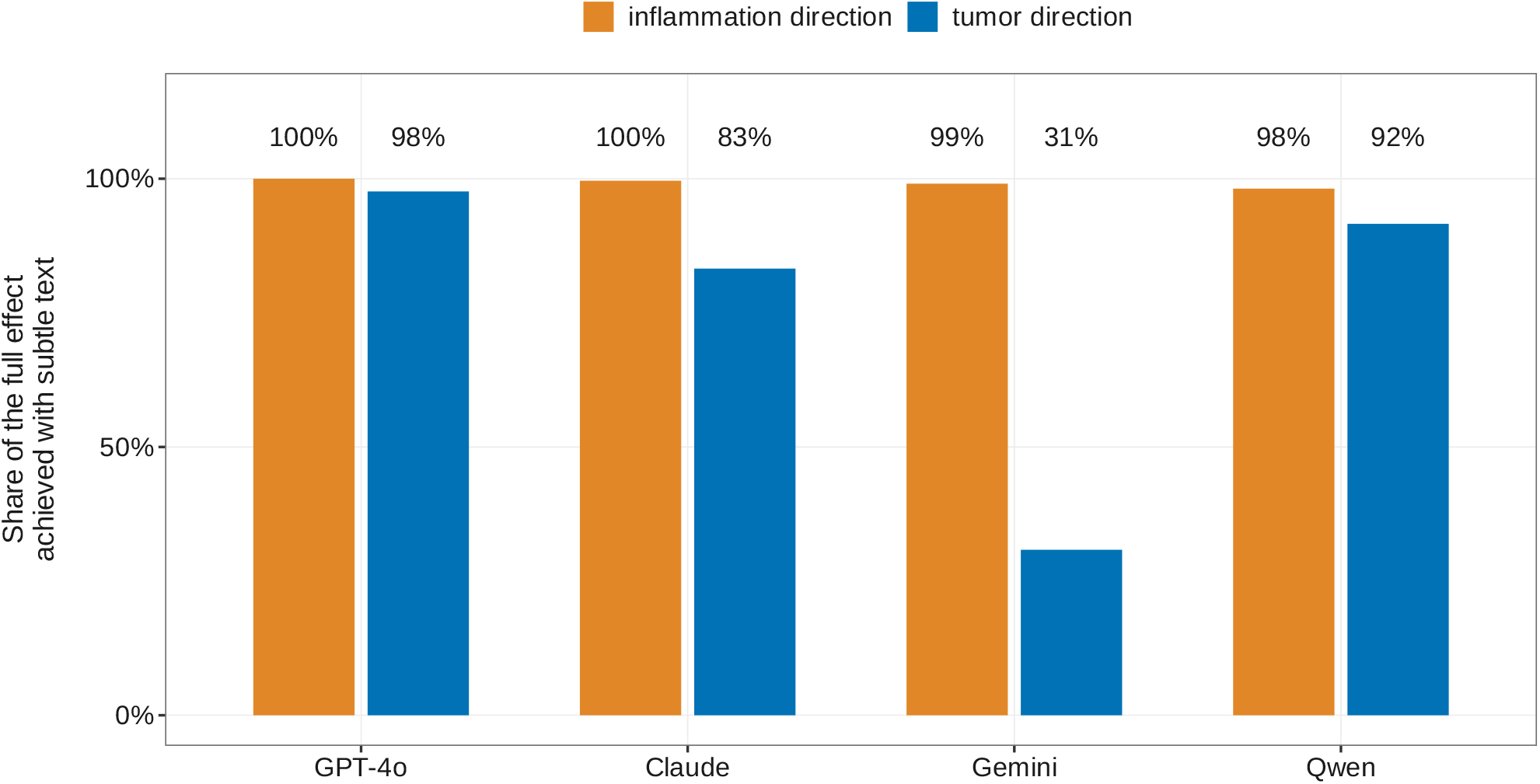
Share of the text effect reached with subtle text. For each model, the change between the explicit inflammatory and the explicit tumor text was set to 100%, and the share already reached with the subtle text is shown for each direction. A value of 100% means that the subtle text alone produced the entire change

### Effect of prompt instructions

Instructing the models to ignore the text reduced the span to a markedly different degree (Fig. 5 and Supplementary Fig. 5). The span fell from 0.79 to 0.04 in Gemini but decreased to a lesser extent in the other models (GPT-4o, 1.00 to 0.58; Claude, 0.97 to 0.48; Qwen, 0.86 to 0.70). The instruction to prioritize the image was weaker throughout (GPT-4o, 0.87; Claude, 0.75; Gemini, 0.31; Qwen, 0.82). In Gemini, the reduction did not appear to result from uniform answers: under this instruction, confirmed tumors were called tumor in 54% to 66% and confirmed abscesses in 18% to 23% of queries, largely irrespective of the text (Supplementary Fig. 5). Qwen appeared least responsive to either instruction.

**Fig 5.**
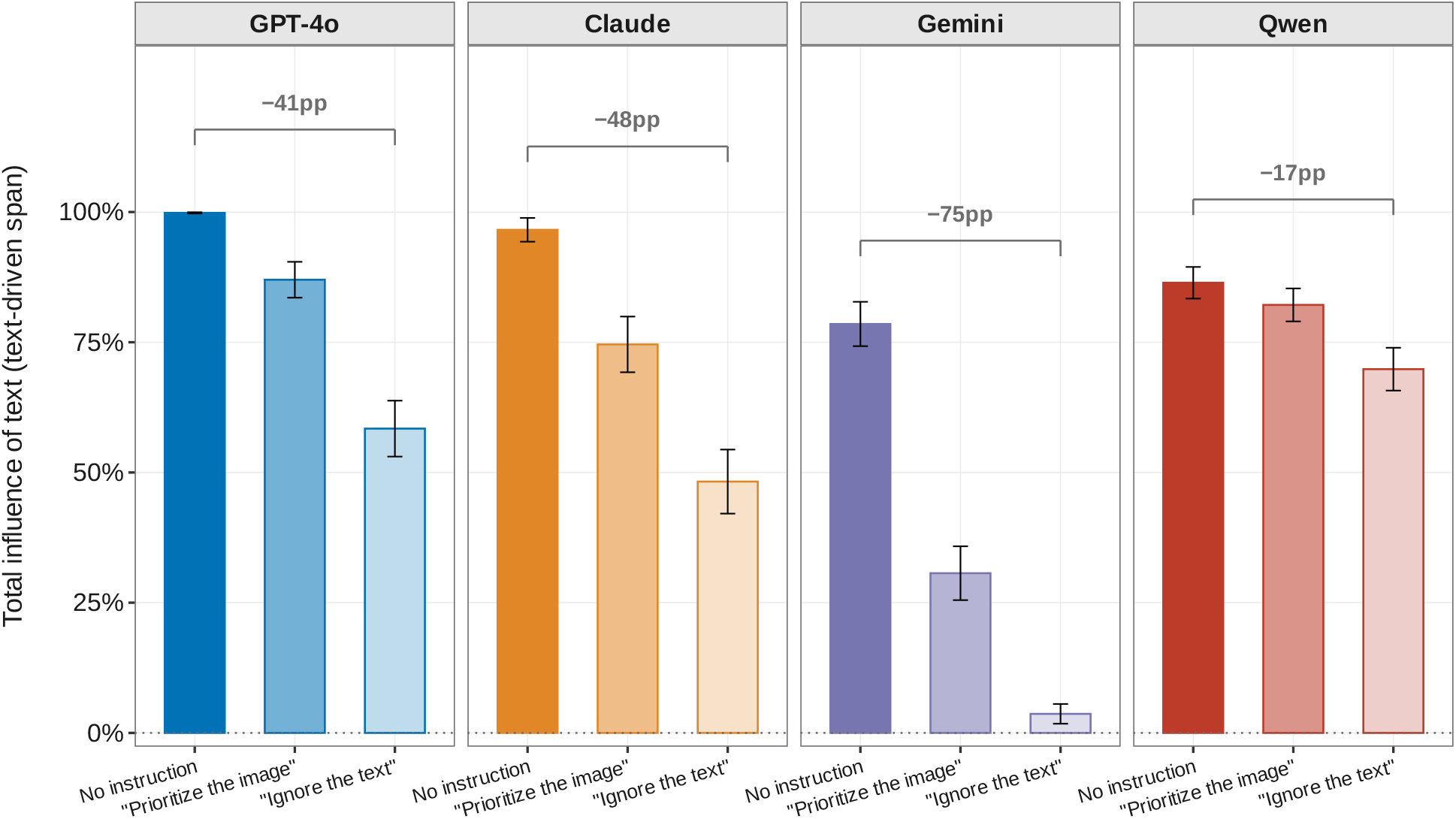
Effect of prompt instructions. Each bar shows the text-driven span under one instruction: none, prioritize the image, or ignore the text. Zero would mean that the text had no influence. Brackets give the reduction in percentage points (pp) from no instruction to the instruction to ignore the text. In Gemini, the separation between confirmed tumors and abscesses was largely preserved under this instruction. Error bars indicate ± 1 standard error

### Consistency

Consistency carried no warning signal: models repeated the same answer nearly as often when their majority answer was wrong as when it was correct (GPT-4o, 0.465 vs. 0.474; Claude, 0.488 vs. 0.497; Gemini, 0.456 vs. 0.474; Qwen, 0.419 vs. 0.453) (Supplementary Fig. 4).

## Discussion

With neutral text, the four multimodal LLMs answered tumor in 45% to 67% of a balanced case set. Suggestive text then moved the answer across almost its entire range, subtle wording was nearly as effective as explicit wording, and instructions to ignore the text reduced the effect only partly and inconsistently. The central finding is therefore not that these models are imperfect classifiers on an ambiguous sequence, but that a single line of clinical text can determine the answer largely independently of the image. Even before any suggestive text was involved, the models appeared to differ in their leaning, with a tendency toward tumor in Claude (specificity for inflammatory lesions, 0.38) and a specificity closer to that of the radiologists in Gemini (0.78). A tumor lean might reflect real-world prevalence and thus a reasonable prior, but the models ranged from 45% to 67% on the identical case set, which argues against a shared prior and for model-specific defaults. Gemini behaved somewhat differently: it separated confirmed tumors and abscesses more clearly than the other models, and when instructed to ignore the text, its answers became largely independent of the text while this separation was largely preserved. This suggests that use of the image and susceptibility to text may be partly distinct properties, which evaluations reporting only one of them, or only accuracy on a balanced set, could miss. Ambiguity of the image warrants uncertainty rather than wholesale adoption of the text, and the radiologists showed that the images still carried diagnostic information. The erroneous calls also carried no warning: models repeated wrong answers nearly as consistently as correct ones, so consistency across repeated queries cannot serve as a confidence signal.

These findings converge with recent evidence that cognitively biased prompts degrade multimodal radiology reasoning [10]. We extend that work in three ways: the text contained no deceptive intent, the images were real cases with reference-standard diagnoses rather than examination items, and the graded five-level design showed how little wording is needed and in which direction the models lean before any text is added. Cognitive biases such as framing and anchoring effects have been documented in clinical LLMs [15, 16], and stigmatizing or authority-laden language can shift model recommendations [17, 18]; our results show the same fragility in an image-based task with ordinary clinical input. That clinical text also influences radiologists is well established. Across 22 studies, clinical history improved diagnostic performance in 15, reduced it in 1, and had no significant effect in 6 [19], and in mammography, misleading history produced small directional changes rather than wholesale shifts [20]. Because radiologists tend to follow confident but incorrect machine suggestions [21, 22], a system that is as certain when wrong as when right is particularly hazardous in routine use. Proposed safeguards include deployment-time guardrails and broader technical and governance controls [23–25], yet a plain instruction to prioritize the image reduced the effect only partly, and an instruction to ignore the text substantially reduced it in only one of four models. Prompt-level safeguards therefore cannot be assumed to generalize and must be validated for each model.

This study has limitations. First, text–image weighting was tested in one disease pair, on a single slice of one sequence, in 60 cases from one institution; whether the magnitude transfers to other diagnoses, modalities, and complete examinations remains to be shown. An unequivocal image could, however, have masked susceptibility to text, which is why the ambiguous sequence was essential to the design. Second, the balanced case mix does not reflect clinical prevalence, and we did not attempt to attribute each model’s leaning to a cause. Third, decoding settings could not be fully harmonized: Claude ran at its provider-default temperature, and output limits differed to accommodate reasoning tokens. Proprietary models also change over time, so the results apply to the versions tested. Fourth, the human reference comprised two readers who received no text, whereas the models received a neutral text because a multimodal query requires some prompt structure; model–reader differences are therefore descriptive, and the readers’ own susceptibility to text could not be quantified. Fifth, the forced-choice design did not permit abstention, so uncertainty could be inferred only from variation across repetitions. A design with an explicit “uncertain” option would show whether the text changes what the model concludes or only how it resolves a binary choice. Future work should examine whether medically pre-trained or fine-tuned multimodal LLMs are similarly susceptible to clinical referral text.

In conclusion, in four current multimodal LLMs, accompanying clinical text largely overrode the image instead of being weighed against it. Subtle wording sufficed, errors could arise in either direction, from false tumor calls to missed tumors, and prompt-based mitigation was inconsistent and model-specific. Because this asymmetry appeared in every model tested, it is unlikely to be a peculiarity of a single model. Clinical use will require model- and task-specific testing of text susceptibility and default behavior, for which reporting frameworks for LLM studies in radiology [26] offer a natural place, and should not presume that instructing a model to prioritize the image is a dependable safeguard.

## Data Availability

All data produced in the present study are available upon reasonable request to the authors.

## Abbreviations

CI: Confidence interval
CLAIM: Checklist for Artificial Intelligence in Medical Imaging
LLM: Large language model
NR1: NR2 Human readers 1 and 2
SD: Standard deviation
SE: Standard error

## Notes

### Competing Interest Statement

The authors of this manuscript declare relationships with the following companies: F.B. reports consulting activities for Hoppr. D.Z. reports serving on the speakers bureaus of Philips Healthcare, Amboss SE, Novaheal by Amboss, and Acandis GmbH, and receiving consulting honoraria from Gaumont. The remaining authors declare no relationships with any companies whose products or services may be related to the subject matter of the article.

### Author Declarations

Use of MRI-Datasets form the Institute for Diagnostic and Interventional Radiology University Hospital Cologne that were available before the initiation of the study. The ethics committee of the University of Cologne reviewed this retrospective single-center study (reference number 24-1252-retro) and raised no objections.

### Funding

- Else Kröner-Fresenius-Stiftung

