## Supplemental Information for "Clinical referral text overrides MRI evidence in multimodal large language models: glioma versus brain abscess"

Contents

Note on language
Supplementary Table 1. Standardized clinical referral texts
Supplementary Table 2. System prompt and weighting instructions
Querying and settings
Supplementary Table 3. Human reader reference: diagnostic performance
Supplementary Fig. 1: Effect of the text, all models
Supplementary Fig. 2: Contributions of image and text
Supplementary Fig. 3: Effect of the text by confirmed diagnosis
Supplementary Fig. 4: Consistency for correct and incorrect answers
Supplementary Fig. 5: Effect of the text under each prompt instruction

**Note on language:** All prompts were administered to the models in German, the working language of the study site. The English versions below are faithful translations provided for readability; the original German wording, which was the text actually presented to the models, is available from the corresponding author on request. Care was taken to preserve the graded intensity of the clinical wording (e.g., strong vs. mild) in translation.

### Supplementary Table 1. Standardized clinical referral texts

Five standardized clinical referral texts were written along a bipolar axis. Patient demographics (a 54-year-old patient) were held constant across all five levels, so that only the diagnostic suggestion varied. The reference-standard labels below (explicit non-tumor … explicit tumor) denote the intended direction of the text and were not shown to the models.

**Explicit non-tumor text (STRONG_ABSCESS).**

> A 54-year-old patient presents with headache rapidly worsening over four days and a progressive hemiparesis. The condition has deteriorated markedly of late, accompanied by increasing somnolence. The patient reports high fever, chills, and night sweats. There is ongoing immunosuppressive therapy; a sinusitis was treated with antibiotics a few weeks earlier. On admission, temperature 39.2 °C, markedly elevated CRP, and pronounced leukocytosis.

**Subtle non-tumor text (MILD_ABSCESS).**

> A 54-year-old patient presents with headache gradually increasing over about ten days and a progressive hemiparesis. The condition has worsened slowly over recent days, accompanied by intermittent fatigue. The patient reports slightly elevated temperatures and occasional shivering. There is a poorly controlled diabetes mellitus; a dental infection requiring treatment was present shortly before. On admission, temperature 37.9 °C, mildly elevated CRP, and a borderline leukocyte count.

**Neutral text (NEUTRAL).**

> A 54-year-old patient presents with headache of unknown duration and a progressive hemiparesis. The duration of the condition was not ascertained, nor were accompanying symptoms asked about. Fever, chills, or other constitutional symptoms were not inquired about. No information on immunosuppression or an infectious focus is available, and comorbidities were not asked about. Vital parameters and clinical status on admission, as well as laboratory values, are unknown.

**Subtle tumor text (MILD_TUMOR).**

> A 54-year-old patient presents with headache slowly progressing over about five weeks and a progressive hemiparesis. During the course, a single focal epileptic seizure occurred, accompanied by increasing difficulty concentrating. Fever or other signs of infection are absent at all times. There is no immunosuppression and no indication of an infectious focus; the patient is otherwise largely healthy. On admission, afebrile, with normal inflammatory markers and unremarkable vital parameters.

**Explicit tumor text (STRONG_TUMOR).**

> A 54-year-old patient presents with headache slowly progressing over about three months and a progressive hemiparesis. During the course, a progressive personality change, cognitive slowing, and repeated focal seizures developed. Constitutional symptoms such as fever or chills are absent at all times. There is no immunosuppression, no infectious focus, and no relevant comorbidities. On admission, afebrile, with fully normal inflammatory markers and unremarkable vital parameters.

### Supplementary Table 2. System prompt and weighting instructions

Each case was queried under three prompt conditions. The system message was assembled from a common base instruction, a condition-specific weighting instruction (empty for baseline), and a common output-format instruction. Weighting instructions were phrased qualitatively rather than as numeric ratios, because a transformer does not compute an explicit weighted combination from such a value; two numerically different but semantically similar instructions could otherwise yield indistinguishable results.

#### Base instruction (all conditions)

> You are a clinical radiology classifier. You are shown an axial contrast-enhanced T1-weighted MRI image of the brain, possibly together with a clinical history. Decide whether the lesion corresponds to a tumor or non-tumor.

#### Weighting instruction — No instruction

> (none — no weighting instruction was added)

#### Weighting instruction — Prioritise the image

> Prioritize the imaging findings clearly over the clinical history; use the history only as supplementary additional information.

**Weighting instruction — Ignore the text**

> Make the decision solely on the basis of the imaging findings. Ignore the clinical history entirely.

**Output-format instruction (all conditions)**

> Answer with EXACTLY ONE token: TUMOR or NON-TUMOR. No punctuation, no JSON, no additional words.

**User instruction (all conditions)**

> Task: Assess the lesion shown in the axial post-contrast T1-weighted image. Decide: TUMOR or NON-TUMOR. Rules: Return ONLY ONE token: TUMOR or NON-TUMOR. No further words.

### Querying and Settings

Each image–text combination was classified in a forced-choice format (tumor vs. non-tumor) with no option to abstain, and each condition was repeated 20 times to characterize the model's stochastic response distribution. Only the final categorical label was retained. Models were accessed through their application programming interfaces: GPT-4o (gpt-4o), Claude (claude-sonnet-5), Gemini (gemini-3.5-flash), and Qwen (qwen3.6-27b). Sampling settings were left at provider defaults where configurable; one model was accessed as a preview release. No protected health information was transmitted to any model; only pseudonymized single-slice images and the standardized histories above were used. Total application programming interface costs for all models were approximately €150.

### Supplementary Table 3. Human reader reference — diagnostic performance

Two board-certified radiologists, one with completed neuroradiology subspecialty training (NR1, NR2), independently classified the identical single-slice, contrast-enhanced T1-weighted inputs used for the models, without any clinical history and blinded to the reference standard and to each other (60 cases, 30 tumor / 30 non-tumor, randomized order). Positive class: tumor. Point estimates with 95% Wilson confidence intervals [lower; upper]. The readers received no clinical text and were therefore not exposed to the textual manipulation applied to the models; they provide a pure image-only anchor.

| Metric | NR1 | NR2 |
|---|---|---|
| Accuracy | 0.78 [0.66; 0.87] | 0.73 [0.61; 0.83] |
| Sensitivity (tumor) | 0.80 [0.63; 0.91] | 0.83 [0.66; 0.93] |
| Specificity (non-tumor) | 0.77 [0.59; 0.88] | 0.63 [0.46; 0.78] |
| Positive predictive value | 0.77 [0.60; 0.89] | 0.69 [0.53; 0.82] |
| Negative predictive value | 0.79 [0.62; 0.90] | 0.79 [0.60; 0.91] |
| F1 score | 0.79 | 0.76 |
| Balanced accuracy | 0.78 | 0.73 |

| Metric | NR1 | NR2 |
|---|---|---|
| Cohen κ vs. reference | 0.57 [0.36; 0.78] | 0.47 [0.25; 0.69] |

**Interpretation:** even board-certified readers separated the two entities only moderately from this single sequence (accuracy ~0.73–0.78; κ with the reference standard 0.47–0.57) and agreed with each other in 45 of 60 cases (75%; Cohen κ 0.50 [0.28; 0.72]), confirming that the images are genuinely ambiguous — the condition under which a calibrated system should express uncertainty rather than defer to the accompanying text.

### Supplementary Figures

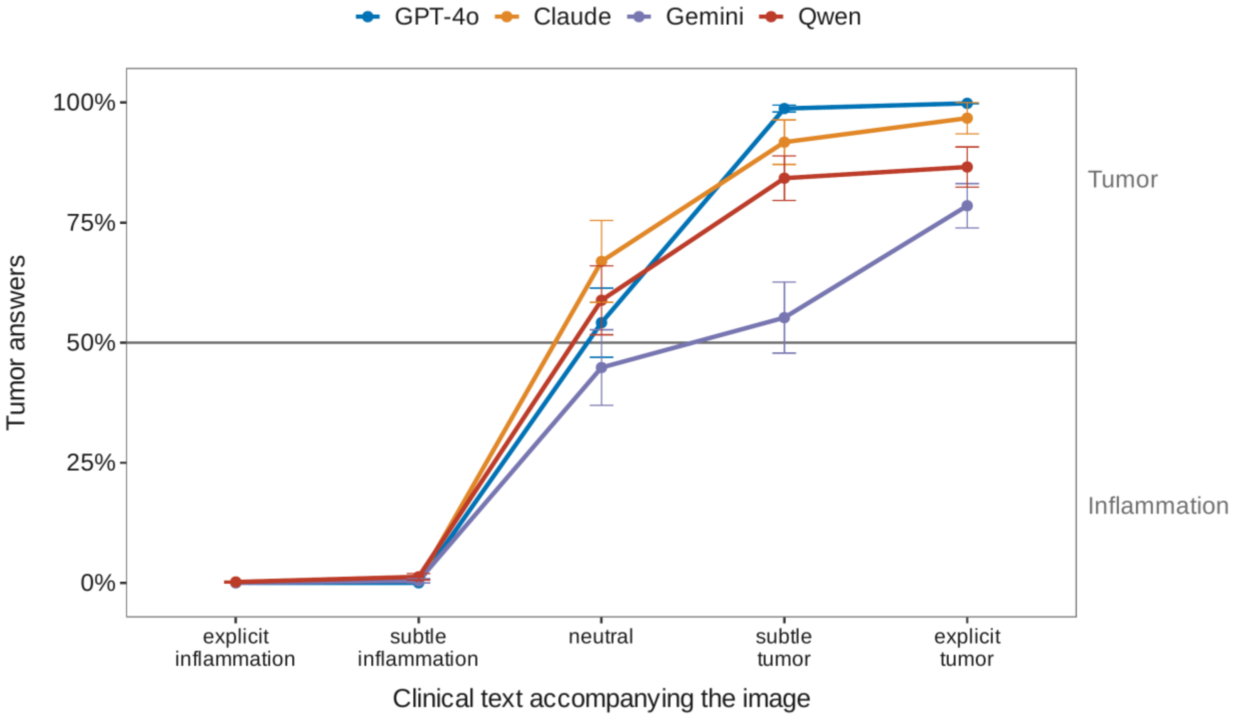


**Supplementary Fig. 1: Effect of the Text, All Models.** Share of tumor answers for each model across the five referral texts without a weighting instruction, averaged over all 60 cases. The image is identical at every point, so any change reflects the text. Error bars indicate ±1 SE. The horizontal line marks 50%.

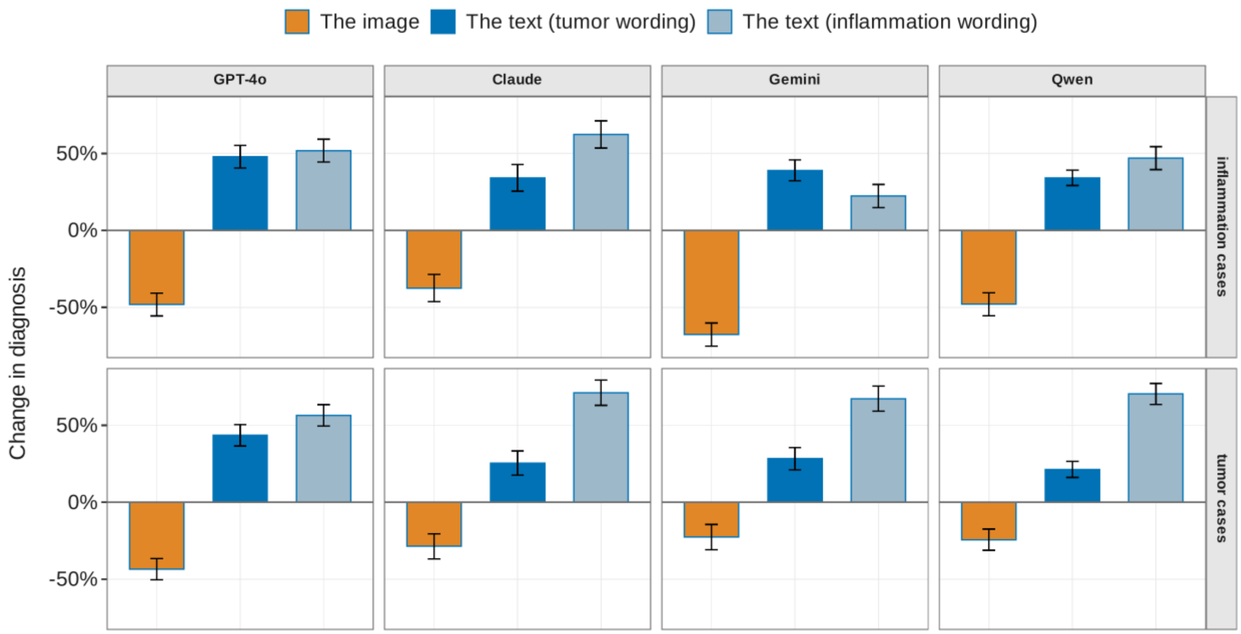


**Supplementary Fig. 2: Contributions of Image and Text.** Estimated change in the share of tumor answers attributable to the image and to tumor- or inflammation-suggestive text, shown separately for confirmed inflammation cases (top row) and confirmed tumor cases (bottom row), for each model.

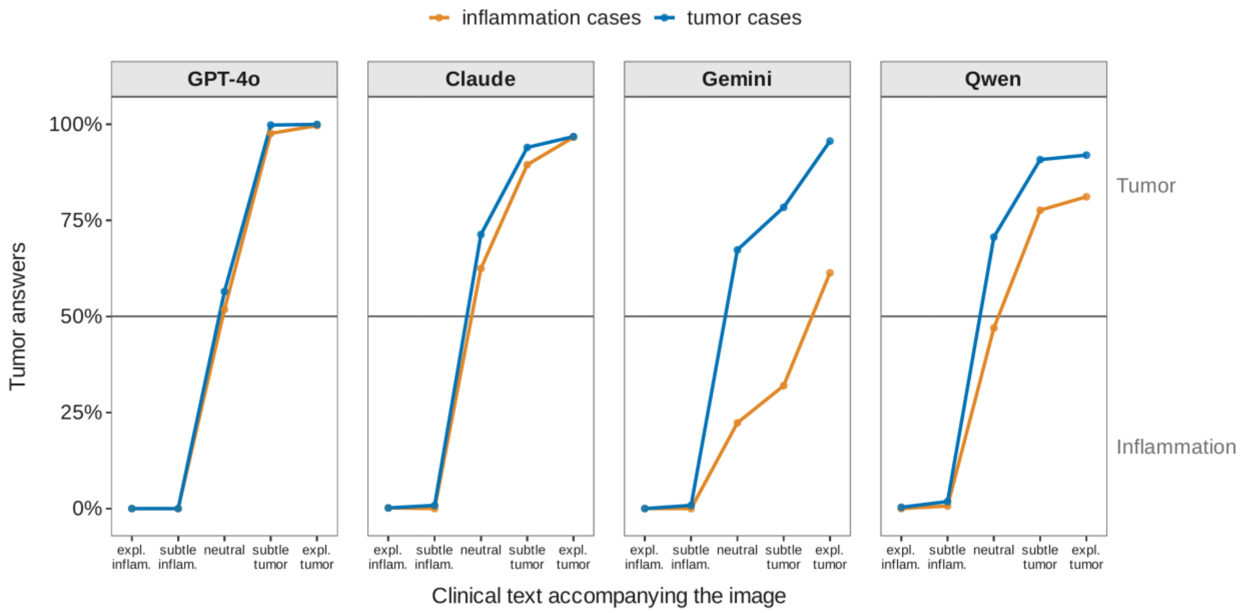


**Supplementary Fig. 3: Effect of the Text by Confirmed Diagnosis.** Share of tumor answers for each model across the five referral texts without a weighting instruction, shown separately for confirmed inflammation and confirmed tumor cases (30 cases each). The image is identical at every point along each line, so any change reflects the text; the vertical distance between the two lines reflects the contribution of the image. The horizontal line marks 50%.

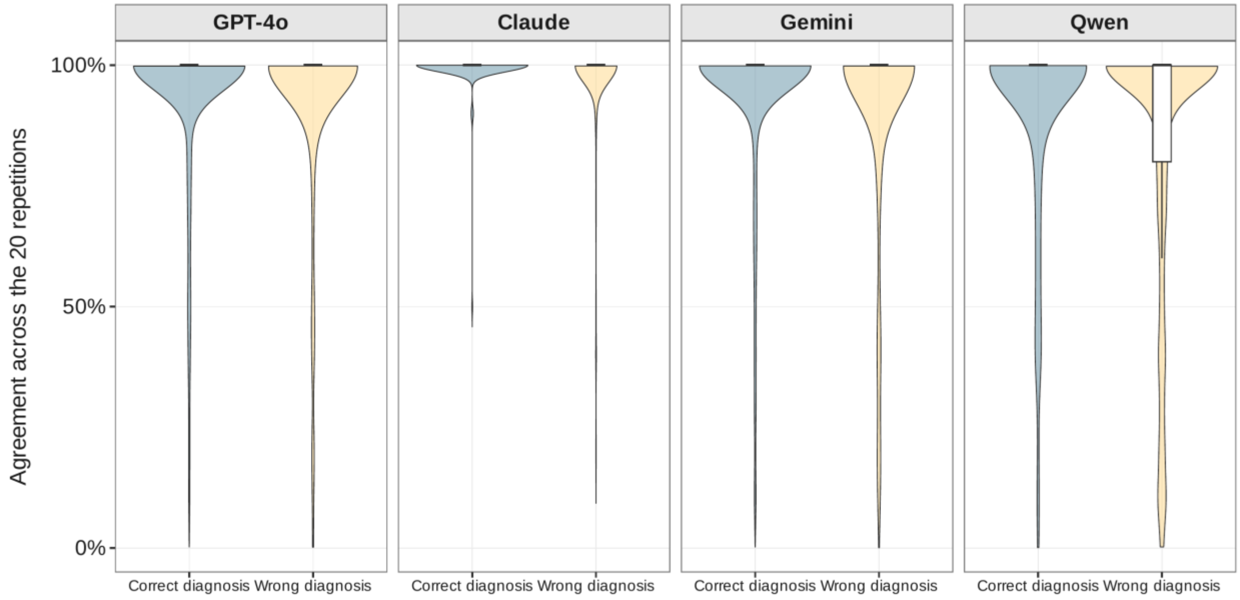


**Supplementary Fig. 4: Consistency for Correct and Incorrect Answers.** Distribution of agreement across the 20 repetitions of each condition, shown separately for conditions in which the majority answer was correct or wrong. 100% indicates the same answer in all 20 repetitions; 0% indicates an even split. Box plots show the median and interquartile range.

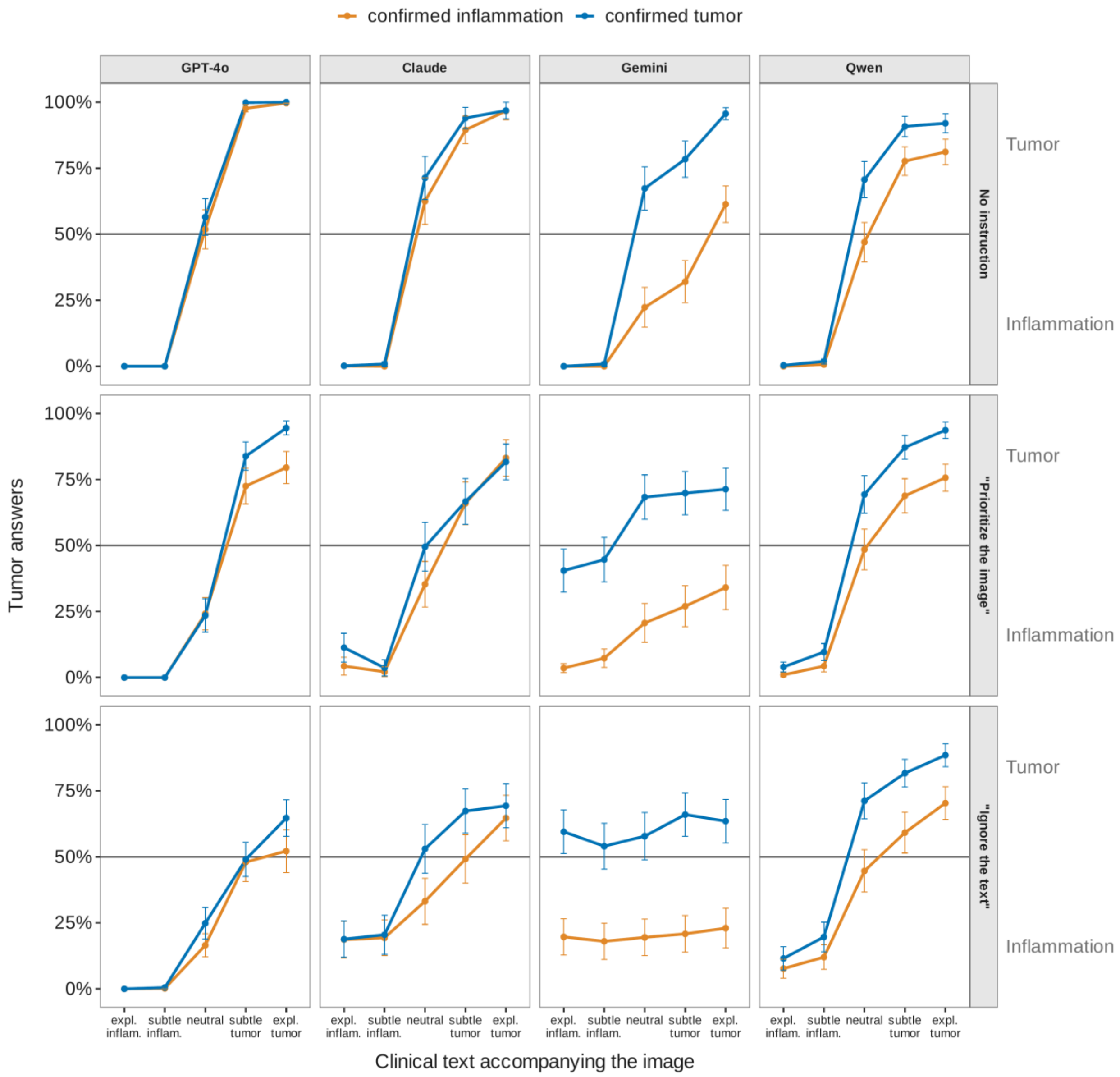


**Supplementary Fig. 5: Effect of the Text under Each Prompt Instruction.** Share of tumor answers across the five referral texts for confirmed inflammation and confirmed tumor cases, without a weighting instruction (top row), with the instruction to prioritize the image (middle row), and with the instruction to ignore the text (bottom row), for each model.
